# A Simple Colorimetric Molecular Detection of Novel Coronavirus (COVID-19), an Essential Diagnostic Tool for Pandemic Screening

**DOI:** 10.1101/2020.04.10.20060293

**Authors:** Pazhanimuthu Annamalai, Madhu Kanta, Pazhanivel Ramu, Baskar Ravi, Kokilavani Veerapandian, Rengarajan Srinivasan

## Abstract

The recent outbreak of the newly emerged novel coronavirus (SARS-CoV-2) presents a big challenge for public health laboratories as virus isolates are not available while there is an increasing evidence that the epidemic is more widespread than initially thought, as well as spreading internationally across borders through travellers does already happen warranting a methodology for the rapid detection of the infection to control SARS-CoV-2. Aim: We intended to develop and deploy a robust and rapid diagnostic methodology using LAMP assay for use in point of care settings to detect SARS-COV-2 infection. Methodology: In the present study, we have developed a validated rapid diagnostic procedure to detect SARS-CoV-2 using LAMP assay, its design relying on isothermal amplification of the nucleic acids of the SARS-CoV-2. Results: The LAMP assay developed detects SARS-CoV-2 infection rapidly with high sensitivity and reliability. The data generated by LAMP assay were comparable and at par with the data generated by real-time PCR method. Conclusion: The present study demonstrates that the LAMP assay developed was a rapid, reliable, sensitive and cost effective method to detect SARS-CoV-2 infection in a point of care as well as in laboratory settings.

## Introduction

Novel coronavirus (SARS-CoV-2) infection is thought to have spread to human populations from the animal kingdom in November or December, 2019, as implied by the phylogeny of genomic sequences obtained from early cases.^1^ The genetic epidemiology implies that from the commencement of December, 2019, when the first cases were retrospectively found out in Wuhan, the increase in infection has been almost entirely driven by human-to-human transmission and not as the result of persistent spillover. There was a colossal transmission of the virus within a matter of weeks in Wuhan, and people in the resulting links of transmission spread infection by national and international travel during the holidays of Chinese new year.

SARS-CoV-2 appears to have different epidemiological characteristics when compared to SARS-CoV-1. SARS-CoV-2 replicates efficiently in the upper respiratory tract and seems to cause less abrupt onset of symptoms, comparable to the conventional human coronaviruses that are a major reason of common colds in the winter season.^2^ Infected individuals, during the prodrome period, produce a large quantity of virus in the upper respiratory tract, are mobile, and carried on routine day-to-day activities, contributing to the spread of infection. SARS-CoV-2 also has affinity for cells in the lower respiratory tract and can replicate there also, causing radiological evidence of lesion in the lower respiratory tract in patients who do not exhibit clinical pneumonia.^1^ There appear to be three key patterns of the clinical route of infection: mild illness with upper respiratory tract presenting symptoms; non-life-threatening pneumonia; and severe pneumonia with acute respiratory distress syndrome (ARDS) that begins with mild symptoms for 7 – 8 days and then advances to rapid deterioration requiring advanced life support.^3^ At present, SARS-CoV-2 appears to spread from person to person by the same mechanism as other common cold or influenza viruses *i.e*., face to face contact with a sneeze or cough or due to contact with secretions of people who are infected. The role of faecal – oral transmission is yet to be established in SARS-CoV-2 but was found to occur during the SARS outbreak.^4^

A possible scenario based on the existing evidence now is that the newly identified SARS-CoV-2, similar to the seasonal influenza virus, is causing mild and self-limiting disease in majority of people who are infected, with severe disease more possibly among aged people or those with comorbidities, such as diabetes, pulmonary disease, and other chronic conditions.

Viruses of the family Coronaviridae have a single-strand, positive-sense RNA genome of 26 – 32 kilo bases in length.^5^ Coronaviruses have been reported in a number of avian hosts,^6, 7^ as well as in a variety of mammals, including camels, bats, mice, dogs, and cats. Novel mammalian coronaviruses are now regularly been identified.^5^ For instance, an HKU2-related coronavirus of bat origin was identified to be responsible for a fatal acute diarrhea syndrome in pigs in the year 2018.^8^ Amongst several corona viruses that are pathogenic to humans, most are associated with mild clinical symptoms,^5^ with two notable exceptions: severe acute respiratory syndrome (SARS) coronavirus (SARS-CoV), a novel beta-coronavirus that emerged in Guangdong, southern China, in November, 2002^9^ and resulted in more than 8000 human infections and 774 deaths in 37 countries during 2002 – 2003;^10^ and Middle East respiratory syndrome (MERS) coronavirus (MERS-CoV), which was first detected in Saudi Arabia in 2012^11^ and was accountable for 2494 laboratory-confirmed cases of infection and 858 casualties since September, 2012, including 38 deaths following a single introduction into South Korea.^12, 13^ In late December, 2019, several patients with viral pneumonia were found to be epidemiologically associated with the Huanan seafood market in Wuhan, in the Hubei province of China, where a number of non-aquatic animals such as birds and rabbits were also on sale before the outbreak. A novel, human-infecting coronavirus,^14, 15^ temporarily named 2019 novel coronavirus (2019-nCoV), was identified with use of next-generation sequencing. As of Jan 28, 2020, China has reported more than 5900 confirmed cases as well as more than 9000 suspected cases of 2019-nCoV infection across 33 Chinese provinces or municipalities, with 106 casualties. Furthermore, 2019-nCoV has now also been reported in Thailand, Japan, South Korea, Malaysia, Singapore, and the USA. People in the health care industry are at high risk of infection, and health-care-associated intensification of transmission is of great concern as is always the case for emerging infections. Most of the infected patients had a high fever and some had dyspnoea, with chest radiographs revealing invasive lesions in both lungs.^1, 16^ Non-pharmaceutical interventions remain vital for the management of SARS-CoV-2 infection as there is no licensed vaccine or coronavirus antiviral available.

At present, for the qualitative detection of the SARS-CoV-2 in nasopharyngeal swabs, alveolar lavage fluid, sputum, and blood samples, reverse transcription-PCR (RT-PCR) kits are being rapidly developed.^17^ Nevertheless, RT-PCR tests require skilled personnel and well-equipped laboratories that restrict this detection method at point-of-care centres and smaller clinics. The increasing number of suspected cases surpasses the capacity of many hospitals, leaving many patients untested thereby challenging the control of spread of the disease. A rapid, point-of-care molecular diagnostics for the SARS-CoV-2 detection is the urgent need of the hour to address the crisis. Therefore, in the present study we have developed a simple closed-tube molecular diagnostic test for SARS-CoV-2 that can be carried out at point-of-care centres and small clinics by minimally trained personnel without the need for sophisticated equipments. In the present study, we report the development of a rapid and reliable molecular detection method to identify the newly emerged coronavirus SARS-CoV-2 by loop-mediated isothermal amplification assay.

## Methods

### Procurement of RNA Clones of Genes of Interest

The RNA Clones of genes of interest, *N* gene, *ORF* gene and the internal control gene, RNAse P were procured as clones from GenScript, USA. The sequences of RNA clones of the genes of interest are as follows:

N Gene: cacccgcaatcctgctaacaatgctgcaatcgtgctacaacttcctcaaggaacaacattgccaaaaggcttca cgcagaagggagcagaggcggcagtcaagcctcttctcgttcctcatcacgtagtcgcaacagttcaagaaattcaactccaggcagcagtaggggaacttctcctgctagaatggctggcaatggcggtgatgctgctcttgctttgctgctgcttgacagattgaaccagcttgagagcaaaatgtctggtaaaggccaacaacaacaaggccaaactgtcactaagaaatctgctgctgaggcttctaagaagcctcggcaaaaacgtactgccactaaagcatacaatgtaacacaagct

ORF Gene

atcgtgttgtctgtactgccgttgccacatagatcatccaaatcctaaaggattttgtgacttaaaaggtaagtatgtacaaatacctacaacttgtgctaatgaccctgtgggttttacacttaaaaacacagtctgtaccgtctgcggtatgtggaaaggttatggctgtagttgtgatcaactccgcgaacccatgcttcagtcagctgatgcacaatcgtttttaaacgggtttgcggtgtaagtgcagcccgtcttacaccgtgcggcacaggcactagtactgatgtcgtatacagggcttttgacatctacaatgataaagtagctggttttgctaaattcctaaaaactaattgttgtcgcttccaagaaaaggacgaagatgacaatttaattgattcttactttgtagttaagagacacactttctctaactaccaacatgaagaaacaatttataatttacttaaggattgtccagctgttgctaaacat

RNaseP Gene atggcggtgtttgcagatttggacctgcgagcgggttctgacctgaaggctctgcgcggacttgtggagacagccgctcaccttggctattcagttgttgctatcaatcatatcgttgactttaaggaaaagaaacaggaaattgaaaaaccagtagctgtttctgaactcttcacaactttgccaattgtacagggaaaatcaagaccaattaaaattttaactagattaacaattattgtctcggatccatctcactgcaatgttttgagagcaacttcttcaagggcccggctctatgatgttgttgcagtttttccaaagacagaaaagctttttcatattgcttgcacacatttagatgtggatttagtctgcataactgtaacagagaaactaccattttacttcaaaagacctcctattaatgtggcgattgaccgaggcctggcttttgaacttgtctatagccctgctatcaaagactccacaatgagaaggtatacaatttccagtgccctcaatttgatgcaaatctgcaaaggaaagaatgtaattatatctagtgctgcagaaaggcctttagaaataagagggccatatgacgtggcaaatctaggcttgctgtttgggctctctgaaagtgacgccaaggctgcggtgtccaccaactgccgagcagcgcttctccatggagaaactagaaaaactgcttttggaattatctctacagtgaagaaacctcggccatcagaaggagatgaagattgtcttccagcttccaagaaagccaagtgtgagggctga

### LAMP Primers

The LAMP primers (Table 1) targeting genes of interest, N and ORF genes and the internal control RNaseP gene were designed using Primer Explorer ver. 5 (http://primerexplorer.jp/lampv5e/index.html). These primers were synthesized by Eurofins Scientific India Pvt. Ltd, Bangalore, India). The primers used for the amplification of the genes of interest and the internal control gene are given in Table 1.

**Table 1:**
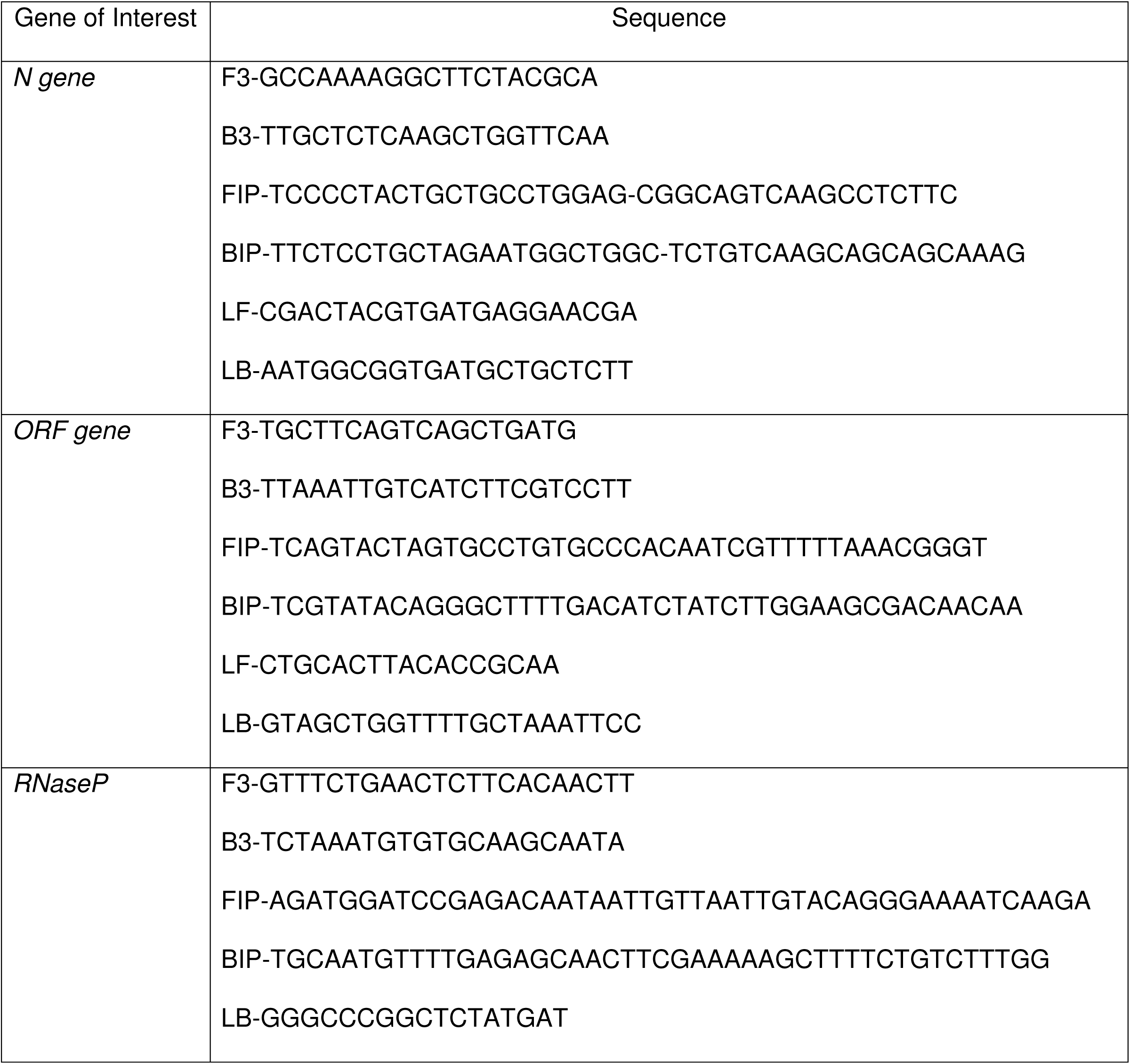
Primers used for LAMP and RT-LAMP

### Colorimetric LAMP Reaction

The LAMP reaction was performed in a total of 25 μl reaction volume using para-cresol dye-based AURA 2 X LAMP master mix (15 µl) that contained each of the four dNTPs (1.4 mM), thermo stable reverse transcriptase (1 µl), AURA *Bst* polymerase (8 U), the primers (1.6 µM FIP and BIP; 0.4 µM LB and LF; 0.2 µM F3 and B3) and 120 µM para-cresol dye. The target RNA at different copy numbers (10^−6^ to 10^0^) was added to the master mix and the final reaction volume was made up to 25 µl with water. NTC had all the reaction components except the target RNA. The reaction was carried out at 65°C for 30 minutes.

### Real-Time LAMP Reaction

The real-time LAMP reaction was performed in a total of 25 μl reaction volume using para-cresol dye-based AURA 2 X LAMP master mix (15 µl) that contained each of the four dNTPs (1.4 mM), thermo stable reverse transcriptase (1 µl), AURA *Bst* polymerase (8 U), the primers (1.6 µM FIP and BIP; 0.4 µM LB and LF; 0.2 µM F3 and B3) and 120 µM para-cresol dye as well as 1 X Evagreen and 1X ROX reference dyes. The target RNA at different copy numbers (10^−6^ – 10^0^) was added to the master mix and the final reaction volume was made up to 25 µl with water. NTC had all the reaction components except the target RNA. The fluorescent signals were observed automatically by MxPro 3000P Real-Time PCR System (Agilent Technologies, New York, USA). The C_t_ values obtained for different copy numbers (10^−6^ – 10^0^) of the N and ORF target RNAs as well as for internal control RNaseP RNA were given in Table 2.

**Table 2:** Cycle Threshold Values

| S. No. | Name of the Target RNA | Copy Number of Target RNA | Cycle Threshold ($C_t$) Value |
| --- | --- | --- | --- |
| 1 | *N* | $10^7$ | 14.13 |
| | | $10^6$ | 15.42 |
| | | $10^5$ | 18.20 |
| | | $10^4$ | 20.16 |
| | | $10^3$ | 24.56 |
| | | $10^2$ | 28.61 |
| | | $10^1$ | 34.36 |
| 2 | *ORF* | $10^7$ | 14.02 |
| | | $10^6$ | 15.62 |
| | | $10^5$ | 17.38 |
| | | $10^4$ | 18.69 |
| | | $10^3$ | 21.99 |
| | | $10^2$ | 28.22 |
| | | $10^1$ | 33.41 |
| 3 | RNaseP | $10^7$ | 21.26 |
| | | $10^6$ | 23.80 |
| | | $10^5$ | 25.13 |
| | | $10^4$ | 26.88 |
| | | $10^3$ | 30.07 |
| | | $10^2$ | 32.62 |
| | | $10^1$ | No CT |

## Results and Discussion

### Real-Time LAMP Detection of SARS-CoV-2

The S-type amplification curves demonstrated a positive result, while the smooth straight line represents the NTC. From the amplification curves, it is evident that the set of primers used for the specific detection of the *N* gene, *ORF* gene and the internal control RNaseP gene in the present study demonstrated increased fluorescence signals in the presence of the templates of the respective genes only and no fluorescence signals was observed for NTC (Fig. 1A, 1B, 1C). The sensitivity was found to be high the viral RNA is detected even up to 10 copy number with good specificity.

**Figure 1.**
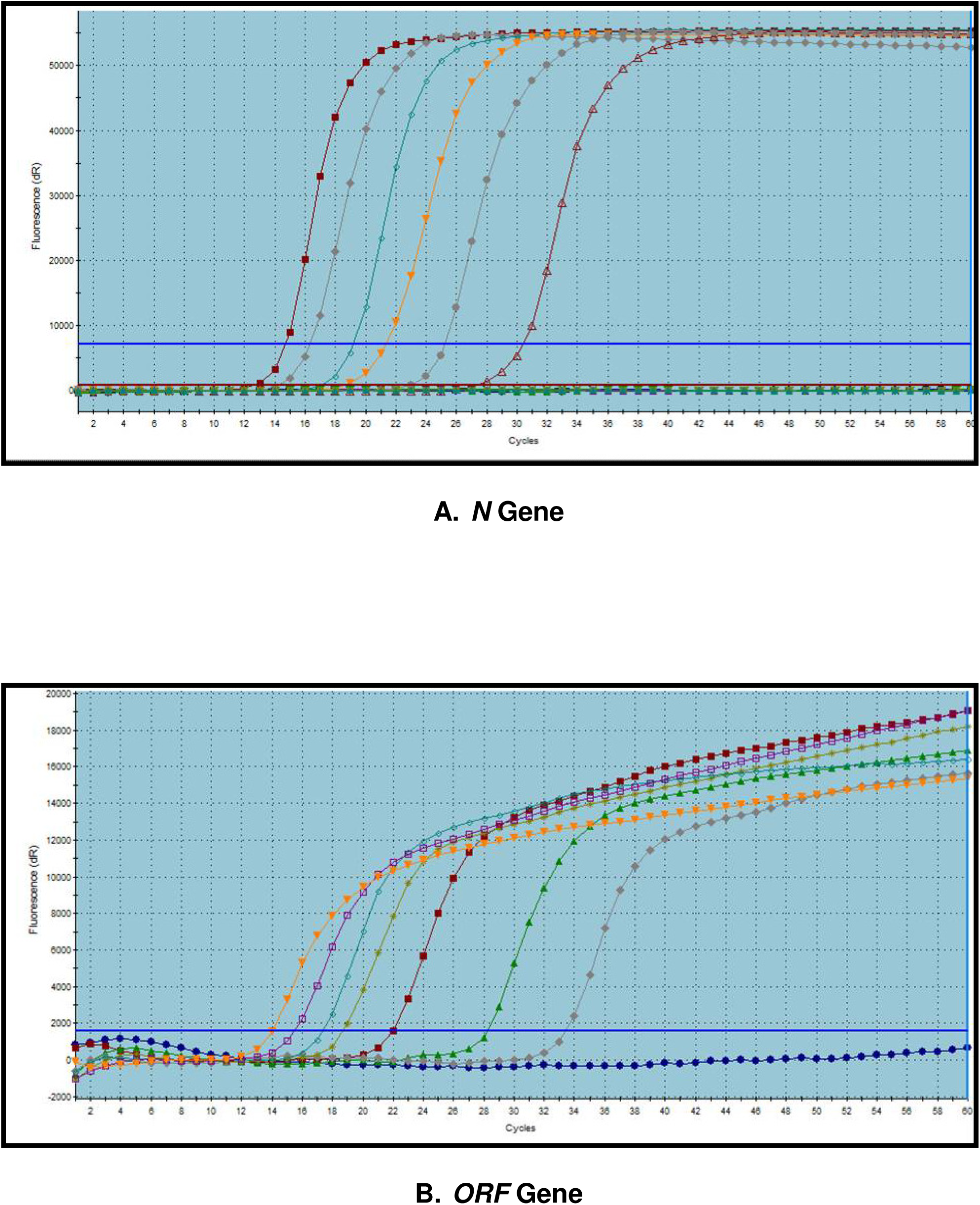
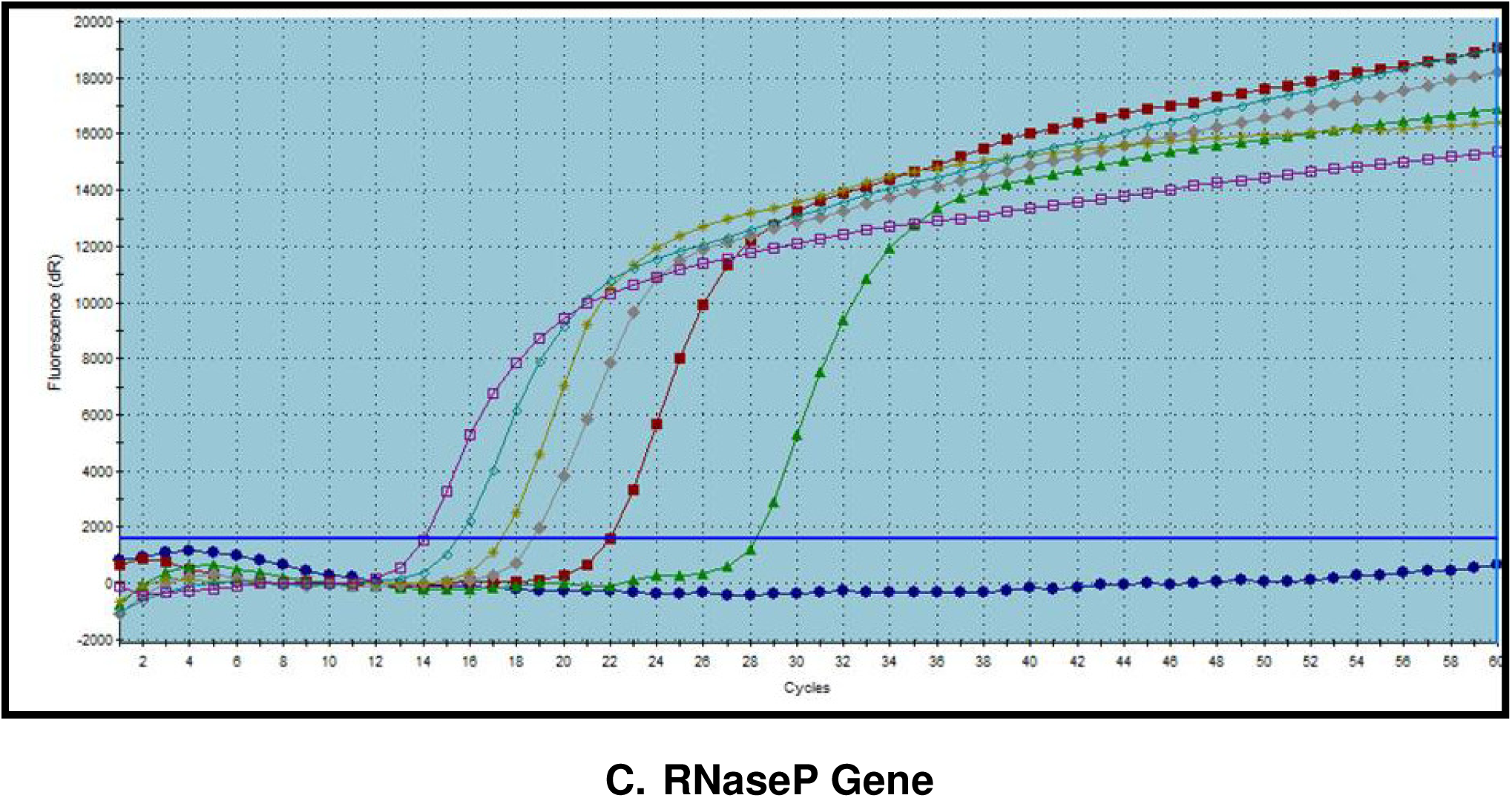

The present study was aimed at developing a LAMP assay specifically for the detection of *N* and *ORF* genes of SARS-CoV-2 (COVID-19) and hence LAMP primers were designed for the amplification of these two genes along with the internal control RNaseP gene using the Primer Explorer (ver. 5) software. The designed primers were validated by performing a real-time LAMP assay with the clones of the *N* and *ORF* gene as well the internal control RNaseP gene and from the amplifications curves generated in the assay, it is observed that the amplicons generated were respective to each of N, ORF and RNaseP genes from the LAMP reaction mix that contained the respective target DNAs and nothing got amplified from the NTC reaction mix thereby demonstrating that the designed primers are the valid ones to use for the detection of SARS-CoV-2 infection in suspected / affected individuals.

### Colorimetric LAMP Detection of SARS-CoV-2

The gradual change in the colour from pink (NTC) to yellow (target DNAs 10^0^) was visually evident as a result of no binding of the target genes’ (N and ORF genes as well as the internal control gene RNaseP) specific primers in NTC LAMP reaction mix to the gradual increased binding of specific primers to the target genes (N and ORF genes as well as the internal control gene RNaseP) of interest in 10-fold serial dilution (10^0^ – 10^−6^) of LAMP reaction mixes (Fig. 2A, 2B, 2C).

**Figure 2.**
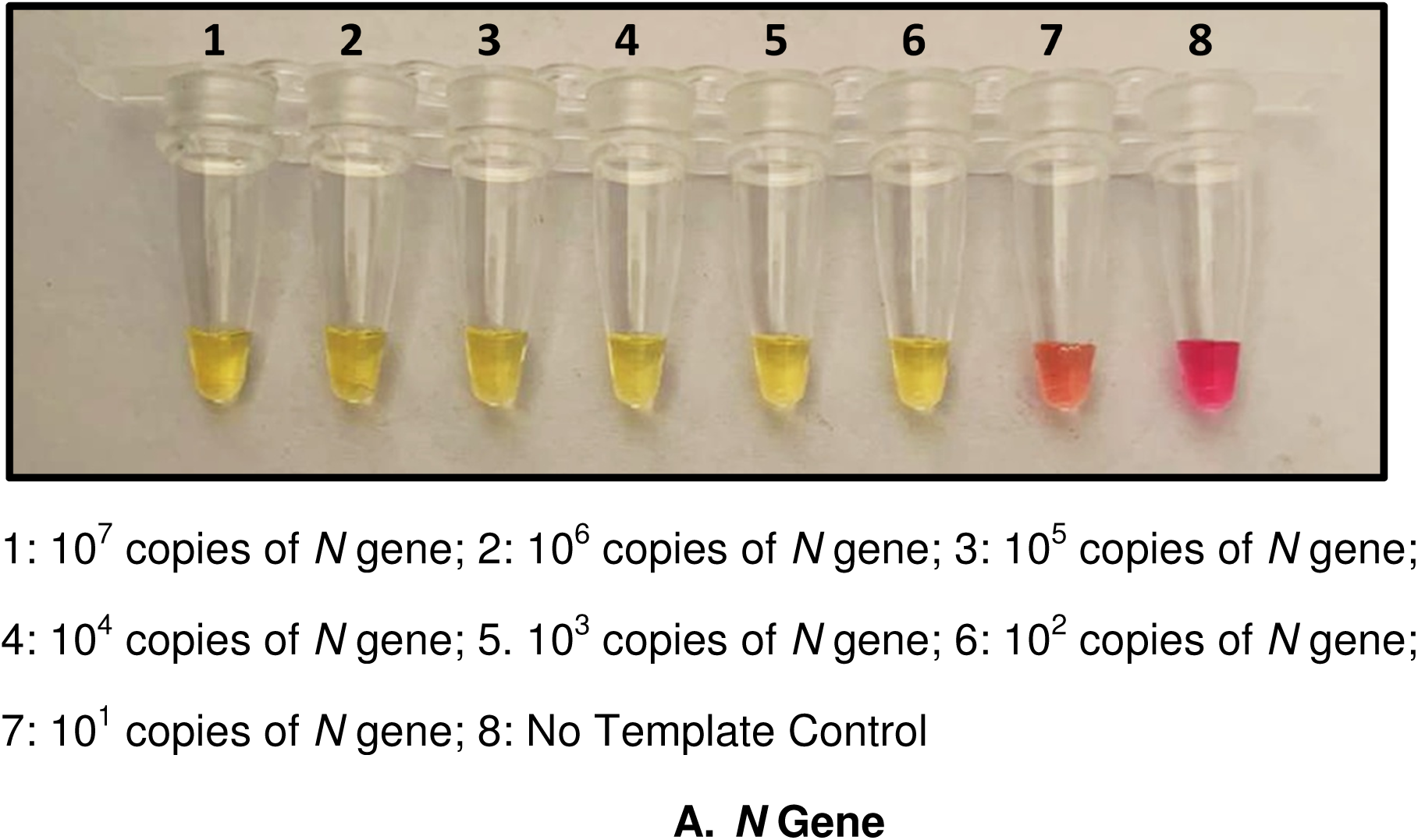
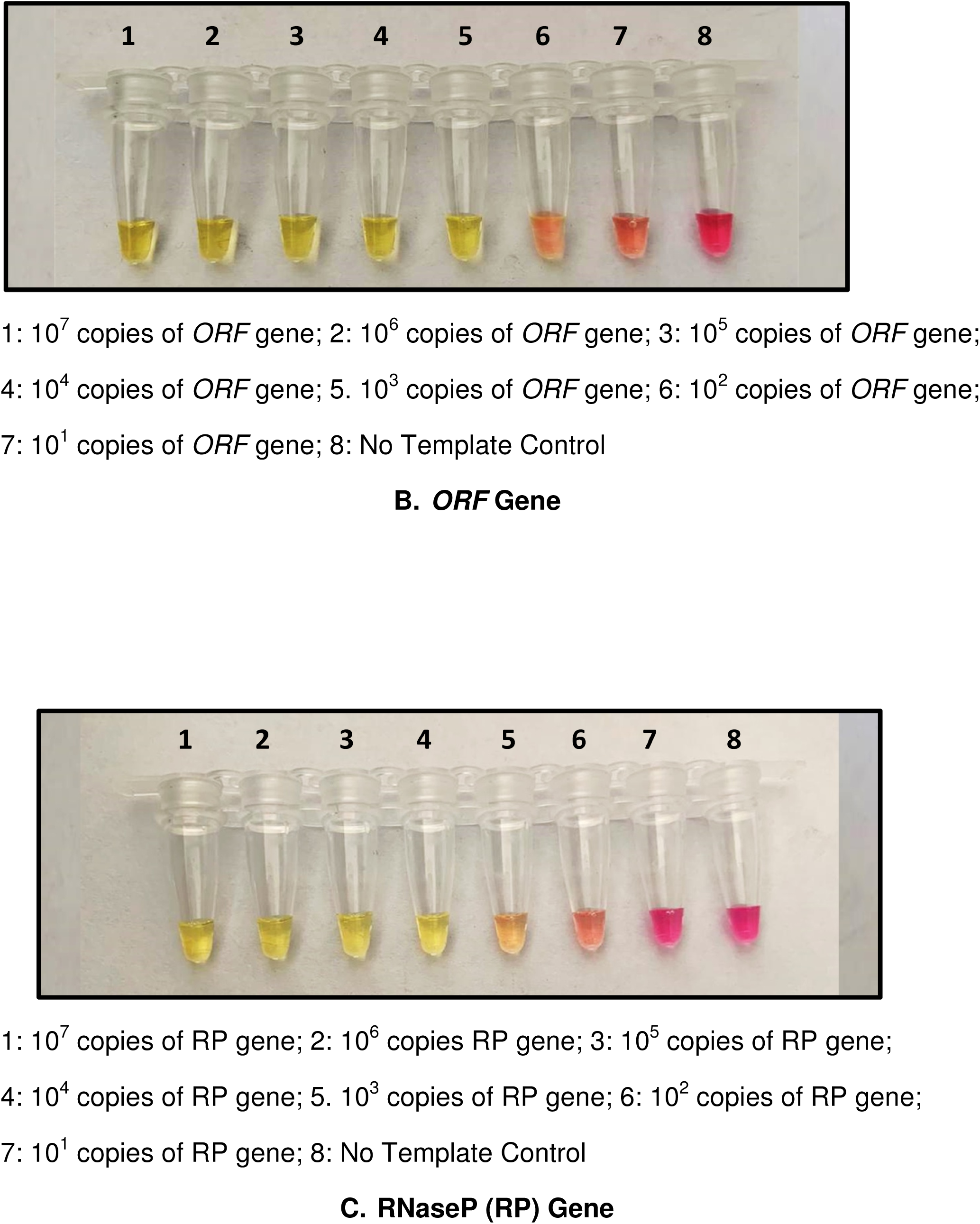

In the current study, at the end of the LAMP assay, we observed a dilution-dependent colour difference (*i.e*., pink to yellow) in the reactions mixes that contained low to high copy numbers of the target DNAs *N* and *ORF* of SARS-CoV-2 as well as the internal control RNaseP gene, thus suggesting that the target DNAs specific primers designed got bound to the specific target DNAs and amplified the same to several magnitude.

## Conclusion

To conclude, the present study has demonstrated that SARS-CoV-2 infection in humans can be detected by LAMP and RT-LAMP assays by targeting the *N* and *ORF* genes of SARS-CoV-2 with high sensitivity and specificity. In addition, our study has also confirmed that LAMP assay is a potential, suitable, cost effective and time saving specific method for the rapid diagnosis of SARS-CoV-2 infection and also aids to a great extent in the screening of this pandemic infection, surveillance as well as combating the spread of SARS-CoV-2 infection at the point of risk.

## Data Availability

All data reported in the manuscript are generated by original research carried out at AURA BIOTECHNOLOGIES PRIVATE LIMITED, CHENNAI, INDIA and the data are available.

